# Cortical networks of parkinsonian gait: a metabolic and functional connectivity study

**DOI:** 10.1101/2023.10.09.23296653

**Authors:** Franziska Pellegrini, Nicoló G. Pozzi, Chiara Palmisano, Giorgio Marotta, Andreas Buck, Stefan Haufe, Ioannis U. Isaias

## Abstract

**Objective:** Locomotion is an automated voluntary movement sustained by coordinated neural synchronization across a distributed brain network. The cerebral cortex is central for adapting the locomotion pattern to the environment and alterations of cortical network dynamics can lead to gait impairments. Gait problems are a common symptom with a still unclear pathophysiology and represent an unmet therapeutical need in Parkinson’s disease. Little is known about the cortical network dynamics of locomotor control in these patients.

**Methods:** We studied the cortical basis of parkinsonian gait by combining metabolic brain imaging with high-density EEG recordings and kinematic measurements performed at rest and during unperturbed overground walking.

**Results:** We found significant changes in functional connectivity between frontal, sensorimotor and visuomotor cortical areas during walking as compared to resting. Specifically, hypokinetic gait was associated with poor information flow from the supplementary motor area (SMA) to precuneus and from calcarine to lingual gyrus as well as high information flow from calcarine to cuneus.

**Interpretation:** Our findings support a role for visuomotor integration processes in PD-related hypokinetic gait and suggest that reinforcing visual information may act as a compensatory strategy to allow SMA-related feedforward locomotor control in PD.

## Introduction

In human locomotion, the synergistic movement of different body parts ensure forward motion while maintaining postural balance under changing environmental conditions. Despite being often considered a highly automated movement, human walking requires a fine integration of multiple brain and spinal processes with converging evidence pointing to a leading role of the brain cortex. In particular, dynamic interaction of distributed cortical areas may be essential for the integration of frontal, sensorimotor and visuomotor information, ^1,2^ which would adapt the stereotypical activity of the spinal central pattern generators to meet the environmental needs ^3,4^. This remarkable motor control can be achieved with synchronous neuronal oscillations, a mean to coordinate the information flow within functionally specialized neural networks (for review ^4–6^). Alterations of these finely tuned dynamics would hamper locomotor control and result in gait impairments.

Gait impairments are a common phenomenon in Parkinson’s disease (PD) and can encompass a wide range of gait disturbances, ranging from hypokinetic gait to episodic gait freezing. In many cases, gait impairments lead to falls, injuries and fear of falling, which can severely affect patient’s quality of life ^7^. While several studies assessed the role of subcortical nuclei ^5,8–10^ in the pathophysiology of gait impairment in PD, recent findings have expanded beyond basal ganglia disfunctions ^10,11^ and suggested a primary role for the brain cortex ^4,12^. Both functional and molecular imaging studies showed that deficits in the integration between frontal and sensorimotor areas might contribute to the disruption of gait automation and favor the development of gait impairments ^4,12^. More recently, studies on cortical visual processing showed that gait impairment correlate with visual perceptual deficits, thus supporting a role for visuomotor processing in the pathophysiology of gait impairment in PD ^4,12^. Yet, a direct assessment of cortical network dynamics during actual locomotion in subjects with PD is still lacking. In particular, it is still unknown how the locomotor information flows within the cortical network in unmedicated PD patients and which integration processes are most relevant for gait under striatal dopamine depletion.

Here, we directly tackle this issue by combining sequential brain metabolic investigations during resting and walking with high-density EEG recordings and kinematic assessments of gait performances in unmedicated PD patients. We leveraged this multimodal approach to describe the cortical network dynamics of locomotion control in PD, detailing the functional roadblocks and rerouting of cortical locomotor information in PD.

## Materials and methods

### Subjects

We studied ten individuals (five females) with idiopathic PD as assessed by means of the UK Brain Bank Criteria. To be included in the study, patients had to be able to walk independently for at least three minutes straight, after pausing all dopaminergic medications for 12 h (meds-off). The presence of any other neurological disease, including cognitive decline (i.e., Mini-Mental State Examination score <27), vestibular disorders or orthopedic impairments were considered exclusion criteria. Two patients were implanted for bilateral subthalamic deep brain stimulation (Medtronic 3389 and Activa PC+S, Medtronic, PLC). Stimulation was turned off two hours prior to the recordings and remained off for all duration of the experiment.

We assessed the motor condition with the Unified Parkinson’s Disease Rating Scale motor part (UPDRS-III) in meds-off and meds-on (i.e., one hour after the intake of the levodopa equivalent daily dose, LEDD) and disease stage with the Hoehn and Yahr (H&Y) scale. We computed the percentage of improvement in the UPDRS-III score due to dopaminergic medication intake, as follows ^10^:

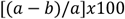

where *a* is the UPDRS-III meds-off score and *b* is the UPDRS-III meds-on score. Demographic and clinical information is listed in Table 1.

**Table 1.** Demographic and clinical data.

| Patient | Sex | Age | Age at onset | LEDD | H&Y | UPDRS-III<br>meds-off | UPDRS-III<br>meds-on | %<br>Improvement |
| --- | --- | --- | --- | --- | --- | --- | --- | --- |
| 1 | F | 60-64 | 50-54 | 680 | 2 | 34 | 24 | 29 |
| 2 | F | 60-64 | 45-49 | 715 | 2 | 20 | 4 | 80 |
| 3 | M | 65-69 | 55-59 | 300 | 1 | 13 | 6 | 54 |
| 4 | M | 60-64 | 50-54 | 610 | 2 | 12 | 5 | 58 |
| 5 | M | 65-69 | 55-59 | 610 | 2 | 30 | 21 | 30 |
| 6 | F | 55-60 | 45-49 | 640 | 1 | 9 | 5 | 44 |
| 7 | M | 55-59 | 50-54 | 900 | 2 | 28 | 5 | 82 |
| 8 | M | 45-49 | 35-39 | 1167 | 3 | 50 | 15 | 70 |
| 9 | F | 65-69 | 60-64 | 440 | 2 | 14 | 4 | 70 |
| 10 | F | 55-59 | 50-54 | 362 | 1 | 18 | 11 | 39 |

Age is expressed in years. F = female and M = male; H&Y = Hoehn and Yahr stage; LEDD = Levodopa Equivalent Daily Dose (expressed in mg); UPDRS-III = Unified Parkinson's Disease Rating Scale motor examination after overnight (>12 h) suspension of all dopaminergic drugs (meds-off) and at 60 min upon receiving 1 to 1.5 times (range 200-300 mg) the levodopa-equivalent of the morning dose (meds-on). % Improvement indicates the percentual amelioration of UPDRS-III score normalized to the UPDRS-III score in meds-off<sup>10</sup>.

The study was approved by the Ethical Committee of the University of Würzburg (n. 36/17 and 103/20) and conformed to the declaration if Helsinki. All participants gave their written informed consent to participation.

### Experimental setup

We investigated brain metabolism and brain functional connectivity at rest and during unperturbed, overground walking (Figure 1). All evaluations were performed in meds-off and took place on two nonconsecutive days in the morning after fasting and pausing all dopaminergic medications for at least 12 h (overnight). Each participant was first investigated at rest. In this condition, participants were asked to comfortably sit on a chair with their eyes open looking at a target on a wall, in a quiet and well illuminated room (i.e., the same used for the forthcoming walking investigation) for 10 minutes before (preconditioning) and for 20 minutes after the injection of [18F]fluorodeoxyglucose (FDG). We recorded cortical electrophysiological activity and monitored body kinematics for 5 minutes before and for 20 minutes after the injection. The PET scan took place 30 minutes after the FDG injection. The same workflow was applied for the studying of gait. Participants walked barefoot at their self-selected speed following a large ellipsoidal path of about 60 m length without perturbations. This setting was chosen to let the subjects walk freely without interruption of the gait pattern. The clinical condition of the participants was monitored by a neurologist trained in movement disorders (I.U.I. and N.G.P.) during the whole period of the study acquisitions. No patient had overt and continuous tremor or other symptoms that might act as a potential confounder of cerebral FDG uptake.

**Figure 1:**
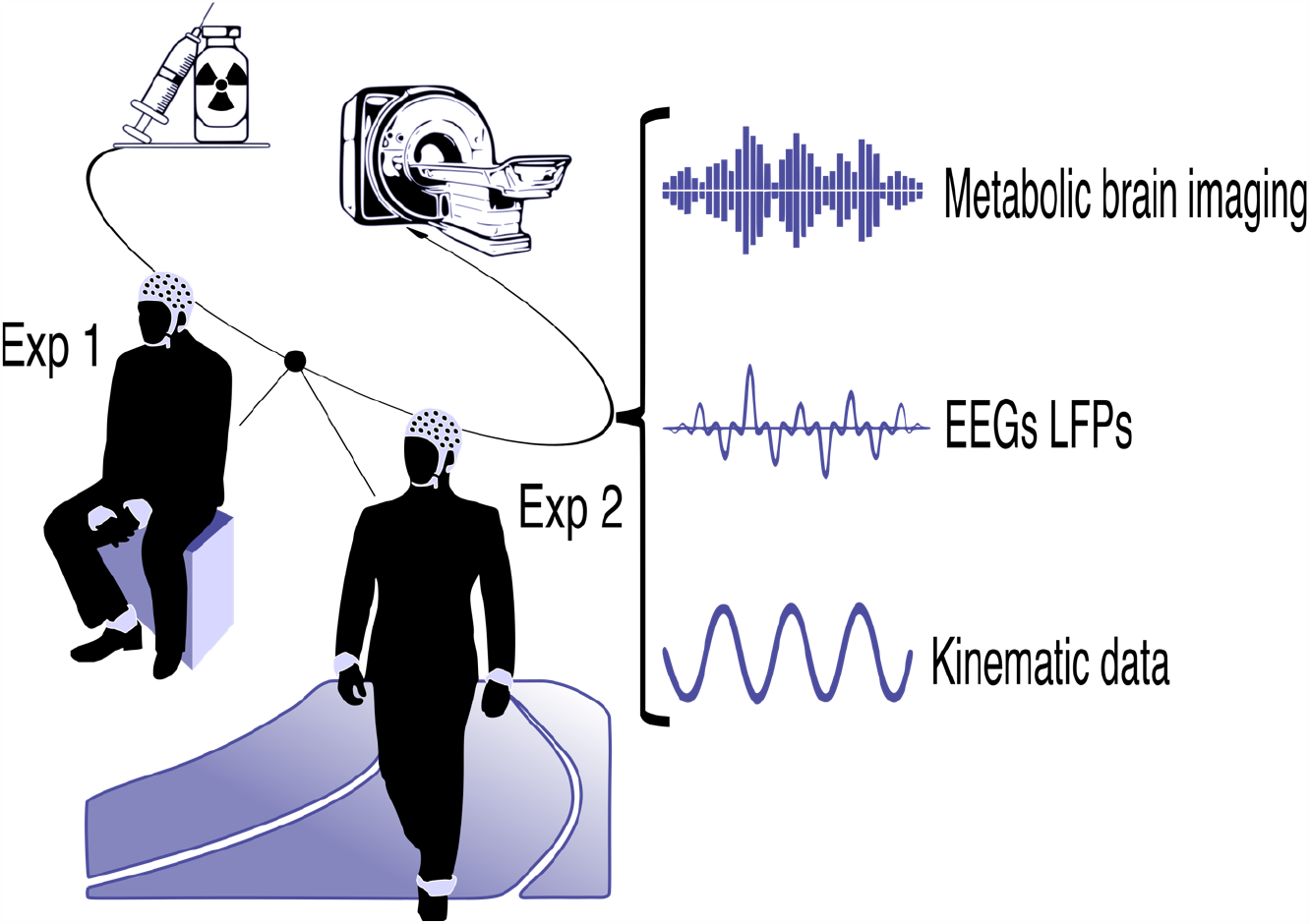
Experimental setup. The simultaneously recorded signals are schematically represented here. Every participant underwent one resting and one walking assessment in two nonconsecutive days in stim-off and meds-off condition (i.e., without subthalamic stimulation and after >12h withdrawal of all dopaminergic medication). Experiments took place always in the morning, participants were fasting >12h. Brain activity (i.e., EEG) and metabolism, and kinematic measurements were assessed simultaneously. Imaging scan took place 30 min after i.v. injection of [18F]fluorodeoxyglucose (FDG). Recordings took place before (preconditioning) and after FDG injection.

**Figure 2:**
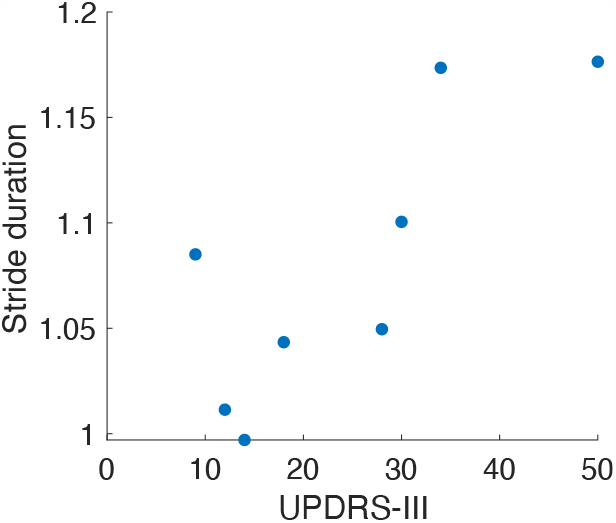
Clinic-Kinematic correlation. Correlation between stride duration and UPDRS-III score (p = 0.017, Pearson’s correlation coefficient = 0.80). Dots represent single subjects.

### Gait kinematics

Throughout the entire walking period, we recorded gait kinematics using three inertial measurement units (IMU, sampling rate of 128 Hz, Opal, APDM, USA) placed on the outer part of the ankles and on the sternum (xiphoid process). In three subjects, we could not acquire IMU recordings due to technical issues. In two of them, we defined gait events with surface electromyography (EMG) as described in ^13^. The second patient (N2) showed artifacts in the EMG signals and was discarded for kinematic analyses. EMG activity was recorded bilaterally from tibialis anterior, soleus, gastrocnemius medialis, gastrocnemius lateralis, and vastus lateralis at a sampling rate of 1000 Hz (FREEEMG, BTS, Italy). IMU, EMG and EEG recordings were synchronized with a transistor-transistor logic (TTL) reference signal and an electrical artifact provided by a transcutaneous electrical nerve stimulation (TENS) device at the beginning and end of each recording session^10^. We detected heel strike and toe-off events from the angular velocity profiles measured with respect to the IMU medio-lateral axis as previously described^13^ and computed: i) swing duration (as the duration between successive toe-off and heel strike events), ii) stride duration (as the time difference between two successive heel strikes), and the swing/stride duration (as ratio of the swing and stride duration).

We further correlate kinematic measurements with clinical scores (i.e., UPDRS-III meds-off) with Pearson correlation.

### Brain metabolic imaging

The execution and analysis of PET scans have been previously described in detail ^14,15^. In brief, all PET/CT scans were performed with a Biograph mCT 64 (Siemens Medical Solutions). For both resting and walking investigations, the PET scan was started 30 min after the injection of 208±16.5 MBq of FDG. PET was performed in 3D mode for 10 min/one bed position using a 400 × 400 matrix with an axial resolution of 2 mm full-width at half-maximum and an in-plane resolution of 4.7 mm. CT scans for attenuation correction were acquired using a low-dose protocol. PET data were reconstructed iteratively (24 subsets, three iterations, Gaussian filtering) using HD reconstruction mode. FDG data processing was performed with statistical parametric mapping software SPM12 (Wellcome Department of Cognitive Neurology, London) implemented in Matlab (MathWorks, Natick, MA, USA). PET images were spatially normalized into the Montreal Neurological Institute standard template (MNI, McGill University, Montreal QC, Canada) using the affine transformation (12 parameters for rigid transformations) of SPM12. Spatially normalized images were then smoothed with a Gaussian filter (FWHM 12 mm) to increase signal-to-noise ratio. Based also on the body weight of each patient, we calculated the standardized uptake value (SUV) of brain regions as defined by the AAL Atlas (PickAtlas Tool, Version 3.0.5 software). After calculating the SUV mean of the white matter in the individual PET, we normalized the SUV of each brain region by the SUV whole mean to remove the effects of the differences in the overall counts. We then computed the change of brain metabolic activity between resting and walking conditions by comparing the relative metabolic changes in a voxel-wise manner. To this aim, we used SPM12 for between-group analysis (i.e., conducting a pairwise t-test for every region). Both metabolism increases and decreases were considered significant for p<0.05 (uncorrected). Significant regions were used as regions of interest (ROIs) in all further analyses (Figure 3).

**Figure 3:**
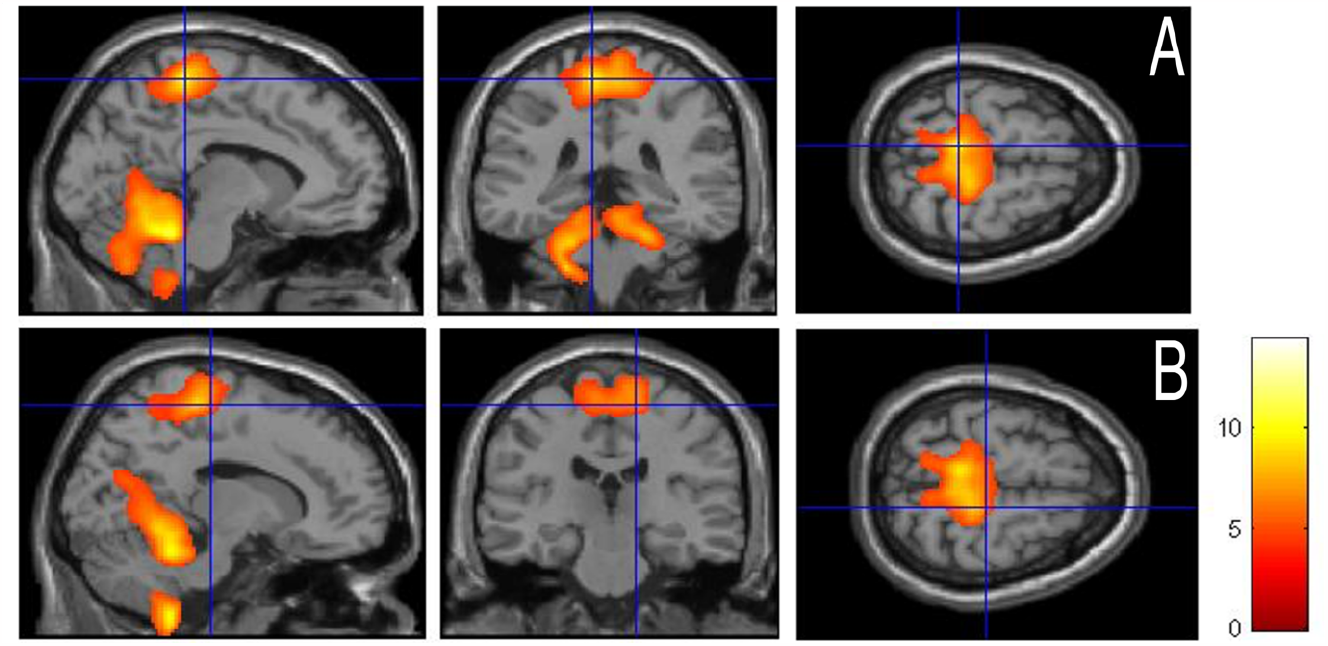
Gait-related metabolic changes (gait vs. resting). The activation clusters are projected onto a standard template provided by SPM in three different views (i.e., sagittal, coronal and axial view from left to right). Top raw (A), crossed blue lines correspond to location (−8, −36, 64), bottom raw (B) crossed blue lines correspond to location (14, −24,62). Brighter colors reflect significantly increased metabolism in the supplementary motor area (SMA), paracentral lobule (BA 4), postcentral gyrus (BA 1-3), superior parietal lobule (BA 5-7), precuneus (BA 7), superior occipital gyrus (BA 19), middle occipital gyrus (BA 18), cuneus (BA 17), calcarine fissure and surrounding cortex (BA 17), lingual gyrus (BA 19). At subcortical level, the thalamus and cerebellum were more active during walking as respect to resting. Pairwise t-test comparison (SPM 12, p<0.001, uncorrected).

### EEG recordings and preprocessing

Neurophysiological recordings were performed using a 64-channel EEG (MOVE, Brain Products) with a sampling rate of 500 Hz. EEG data were resampled to a sampling rate of 125 Hz, zero-phase lowpass-filtered at 45 Hz, and highpass-filtered at 1 Hz (zero-phase forward and reverse second-order digital band-pass Butterworth filter). Line noise was eliminated using a zero-phase bandstop filter with stop band between 47 and 53 Hz. For the walking trials, we discarded the EEG activity in the time-period before the first heel strike and after the last toe-off event as not related to walking. Artifactual channels and segments were identified using automated artifact rejection^16^. We manually detected remaining channels with artifacts by inspecting topographic plots of the alpha power (8-13 Hz). Artifactual channels were replaced with spherical spline interpolation on the scalp. Artifactual segments were removed for independent component analysis (ICA) but were preserved for subsequent analyses to grant for the continuity of the data. To remove eye movements from the data, we regressed out the time series of the two electrodes closest to the eyes (FP1 and FP2) from the time series of all other electrodes. As a further cleaning step, we applied artifact subspace reconstruction ^16^. Then we conducted ICA using runica ^17^ and visually detected and rejected eye, heart, and muscle components. The EEG data were then re-referenced to a common-average reference. Due to severe artefact contamination, we excluded one subject from the cortical analysis (N3).

### Head modeling and source localization

For all but one participant (N9), we constructed individual volume conductor models based on structural MRI data to account for their different head anatomies. In N9, we applied the ICBM-152 template ^18^, as the MRI was unavailable. Where available, lead fields were constructed based on exact channel positions. First, the MRI data were segmented into white matter and grey matter (WM and GM). To this end, we used the recon-all command of the freesurfer software (version 1.0) ^19,20^ to obtain triangularized surface meshes for the WM-GM and GM-CSF interfaces. Based on these segmentations, we reconstructed the outline of the brain. We constructed personalized head models with the boundary element method (BEM) ^21,22^ with three layers (scalp, outer skull, inner skull) using OpenMEEG ^21^ and Brainstorm^23^. MRI and EEG channel positions were co-registered using fiducial points at the nasion, left ear and right ear. The source model consisted of 2000 locations evenly spaced and placed in the center of the gray matter (half-way between the GM-cerebrospinal fluid and WM-GM interfaces). From the individual head model and the cleaned EEG data, we calculated a personalized linear inverse filter (linearly-constrained minimum variance spatial filtering, LCMV) ^24^ to project the EEG channel time series to the source locations. Note that we constructed LCMV filters on the cleaned and filtered times series of every session separately. The LCMV projection yields an estimate of the 3D primary current of an assumed dipolar electrical source, representing the combined activity of a large group of neurons, separately for each source location. On the source level, we labelled every source as being part of one of the regions of the AAL atlas (PickAtlas Tool, Version 3.0.5 software) ^25^.

### Spectral power and directed functional connectivity estimation

We assessed spectral power and directed functional connectivity (FC) within and between AAL regions using a recently validated pipeline ^26^. To avoid a bias due to different trial lengths in the FC measures ^27^, we cut every trial into intervals of one minute length. After cutting, data intervals of under one minute length were discarded from further analysis. Power and FC were then calculated for every interval individually and treated as different samples for linear mixed-effects modeling (LMEs; see also Statistical Analysis Section). To calculate power at every source location, we used the Welch method (2 s non-overlapping epochs, resulting in a 0.5 Hz frequency resolution), and averaged power across sources and 3D dipole orientations within the AAL regions to obtain one power estimate per region. FC was calculated in three steps ^26^. First, we applied dimensionality reduction to the time series of all voxels of each ROI. To this end, we performed a principal component analysis (PCA) and kept the first principal component of every ROI for further processing. Second, we calculated time-reversed spectral Granger causality (TRGC) ^28^ as measure for directed FC between all ROI pairs. Compared to conventional measures of directed FC, TRGC offers empirically and theoretically proven robustness to artifacts of source mixing due to volume conduction in the head or spatial leakage in inverse solutions ^28–30^. Finally, power and FC scores were averaged within two personalized frequency bands of interest (BOIs): a broad low-frequency band containing both theta and alpha and the individual beta band. To derive the borders of the individual frequency bands, we first estimated the individual alpha frequency (IAF) as the alpha peak location on the 1/f-corrected power spectra ^31^. According to ^32^, we defined the theta/alpha band to be between 0.4 * *IAF* and 1.2 * *IAF*, and the beta band to be between 1.2 * *IAF* and 30 Hz. Averaged powers were log-transformed prior to subsequent statistical analysis. Subsequently reported results thus always refer to log power.

### Comparison of spectral power and functional connectivity between resting and walking

To compare power and FC between the resting and walking condition, we calculated an LME for every BOI and ROI or, in case of FC, ROI pair. Specifically, power and FC were calculated for 1min-intervals (see above). These intervals were then presented as samples in the LME. To control for subject- and hemisphere-related biases, subject ID and hemisphere were included as a random and fixed factor, respectively. To correct for multiple comparisons, we used the false discovery rate (FDR) method ^33^. FDR correction was done separately for the analysis of changes between resting and walking, and for the analysis of the relationship between gait kinematics and EEG. Note that we only calculated FC for the lower triangular matrix (see Figure 4 and Figure 5), since the values of the upper triangular matrix are redundant: TRGC_A->B_ = −TRGC_B->A_ (i.e., due to the anti-symmetry of the TRGC measure, any increases in the information flow from a region A to a region B can also be interpreted as a decreased in flow from region B to region A). FDR correction was also applied only on one direction of information flow due to the anti-symmetry.

**Figure 4.**
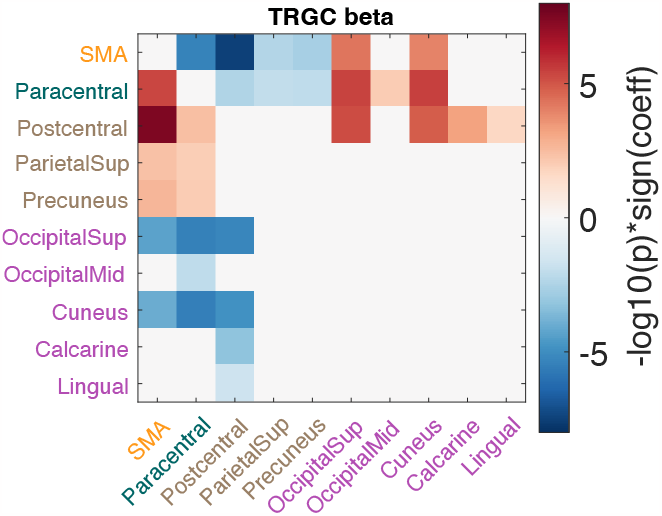
Comparison of EEG markers between resting and walking conditions in the beta band. Changes in directed functional connectivity were estimated by time-reversed Granger causality (TRGC). Red colors either indicate more information flow from the ROI on the Y-axis to the ROI on the X-axis in walking compared to resting, or less information flow from the ROI shown on the X-axis to the ROI shown on the Y-axis. We show log_10_ p-values, thus values below −2 and above 2 would indicate significance at alpha level 0.01. Signed log_10_ p-values that did not survive FDR correction are set to zero.

**Figure 5.**
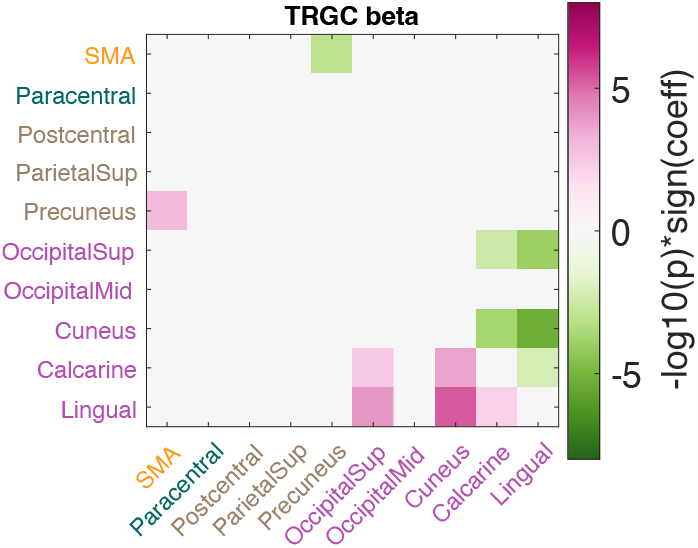
Correlation between stride duration and directed functional connectivity during walking. Pink colors indicate a positive correlation between stride duration and the net information flow from a region plotted on the Y-axis to a region plotted on the X-axis, while green colors indicate a negative correlation. We show log_10_ p-values, thus values below −2 and above 2 would indicate significance at alpha level 0.01. Signed log_10_ p-values that were not significant after FDR correction are set to zero.

## Results

### Clinical and kinematic data

Detailed demographic and clinical data are listed in Table 1. The average age of the participants was 60±7 years and the average disease duration was 9±7 years. The average LEDD at the time of the experiment was 642±256 mg/day. The average UPDRS-III score in meds-off state was 23±13 points and 10±7 points in meds-on (at 60 min upon receiving 1 to 1.5 times the levodopa-equivalent of the morning dose). The average improvement was 56% (range 29% to 71%).

Kinematic measurements of the lower limbs are listed in Table 2. We showed a positive Pearson correlation between UPDRS-III score and stride duration (*p*_*stride-UPRDS*_ = 0.017, coeff_stride-UPDRS_ = 0.80, Figure 2), but not with swing duration and swing/stride duration (*p*_*swing-UPDRS*_ = 0.091, *p*_*swing/stride-UPDRS*_ = 0.98). Therefore, we focused only on stride duration for subsequent analysis.

**Table 2.** Kinematic data.

Kinematic data.
| Patient | Swing duration<br>seconds | Stride duration<br>seconds | Swing duration/Stride duration |
| --- | --- | --- | --- |
| 1 | 0.56 | 1.17 | 0.48 |
| 4 | 0.46 | 1.01 | 0.45 |
| 5 | 0.54 | 1.10 | 0.49 |
| 6 | 0.49 | 1.09 | 0.46 |
| 7 | 0.50 | 1.05 | 0.47 |
| 8 | 0.52 | 1.18 | 0.45 |
| 9 | 0.50 | 1.00 | 0.50 |
| 10 | 0.45 | 1.04 | 0.43 |

The swing duration represent the duration between successive toe-off and heel strike events and the stride duration the time difference between two successive heel strikes; both are expressed in seconds. We calculated also the swing/stride duration, which is the ratio of the swing and stride duration.

### Gait-related metabolic brain changes

We showed a gait-related brain metabolic activation pattern comprising the frontal, parietal and occipital cortical areas. Specifically, the following cortical areas showed an increased FDG uptake during walking as compared to resting: supplementary motor area (SMA), paracentral lobule, postcentral gyrus, superior parietal lobule, precuneus, superior occipital gyrus, middle occipital gyrus, cuneus, calcarine fissure and surrounding cortex, lingual gyrus. At subcortical level, we observed activation of the thalamus and cerebellum during walking. These two areas have not been studied further because it is still debated whether they can be reliably recorded with EEG. Of relevance, we did not find any brain region with reduced FDG uptake during walking as compared to resting. The metabolic brain imaging patterns are shown in Figure 3.

### EEG power and functional connectivity during walking

We observed an increase in power for both frequency bands (i.e., theta-alpha and beta) and over all ROIs in walking as compared to resting, thus matching the brain metabolic changes (supplementary figure S1). Then, we estimated directed functional connectivity (i.e., TRGC) and showed changes between frontal, parietal and occipital areas in PD patients during walking with respect to resting (Figure 4). In the beta band, during walking we showed reduced information flow from the SMA to motor areas corresponding to the lower limb representation in the motor and sensory homunculus (i.e., paracentral) and sensory areas (i.e., postcentral, parietal sup. and precuneus). In contrast, functional connectivity from SMA to visuomotor areas (i.e., cuneus and superior occipital gyrus) was increased during walking compared to resting. The paracentral lobule showed an increased information flow to the SMA and to visuomotor areas, but a reduced connectivity to sensory areas. Sensory areas showed increased functional connectivity with SMA, motor (i.e., the paracentral lobule) and the visuomotor areas during walking with respect to resting. Finally, the visuomotor areas presented a marked reduction in functional connectivity with SMA, motor and sensory areas. Of note, the medial occipital lobule and the lingual gyrus showed the poorest connectivity reduction. The same pattern of functional connectivity changes described for the beta band was shown overall also for the theta-alpha bands (supplementary figure S2).

### Correlation of cortical network dynamics changes and gait kinematics

To test the relevance of the changes in cortical network dynamics, we investigated their correlation with gait performances as assessed by means of kinematic measurements.

We observed an overall negative relationship between stride duration and EEG power, thus supporting a functional role for the selected areas in gait control.

We then observed that only changes in the beta frequency band correlated with the motor performance. Furthermore, we observed that only the connectivity of SMA, precuneus and visuomotor areas was related to gait kinematics. Specifically, we found that the lower the information flow from SMA to precuneus, the higher the stride duration, and thus, the slowest was the gait (hypokinetic gait) (Figure 5). Interestingly, despite not having shown changes in connectivity within visuomotor areas between walking and resting, we noticed that hypokinetic gait was related to poor information flow from visuomotor areas (i.e., occipital superior and cuneus) to lingual gyrus and high information flow calcarine and lingual gyrus to cuneus.

## Discussion

Understanding the neural network dynamics underpinning human locomotion is among the greatest challenges in neuroscience and can lay the foundation for novel therapeutic approaches for PD gait impairments^12^. Recent evidence points to a central role for a distributed network of cortical areas in the locomotor control in subjects with PD ^5,10,34^. In particular, dynamic synchronization of neural oscillations across functionally connected cortical areas would support motor, sensorimotor and visuomotor processes that sustain an active control of locomotion ^4,6^. We leveraged brain metabolic investigation during gait to assess locomotor networks dynamics and estimated the cortical functional connectivity changes that are relevant for gait in subjects with PD (Figure 4). We show a predominant involvement of the SMA, sensory and visuomotor brain areas. We further identified three main functional connectivity patterns that correlate with gait performance (Figure 5) and suggest a putative roadmap for the flow of locomotor information in PD (Figure 6).

**Figure 6.**
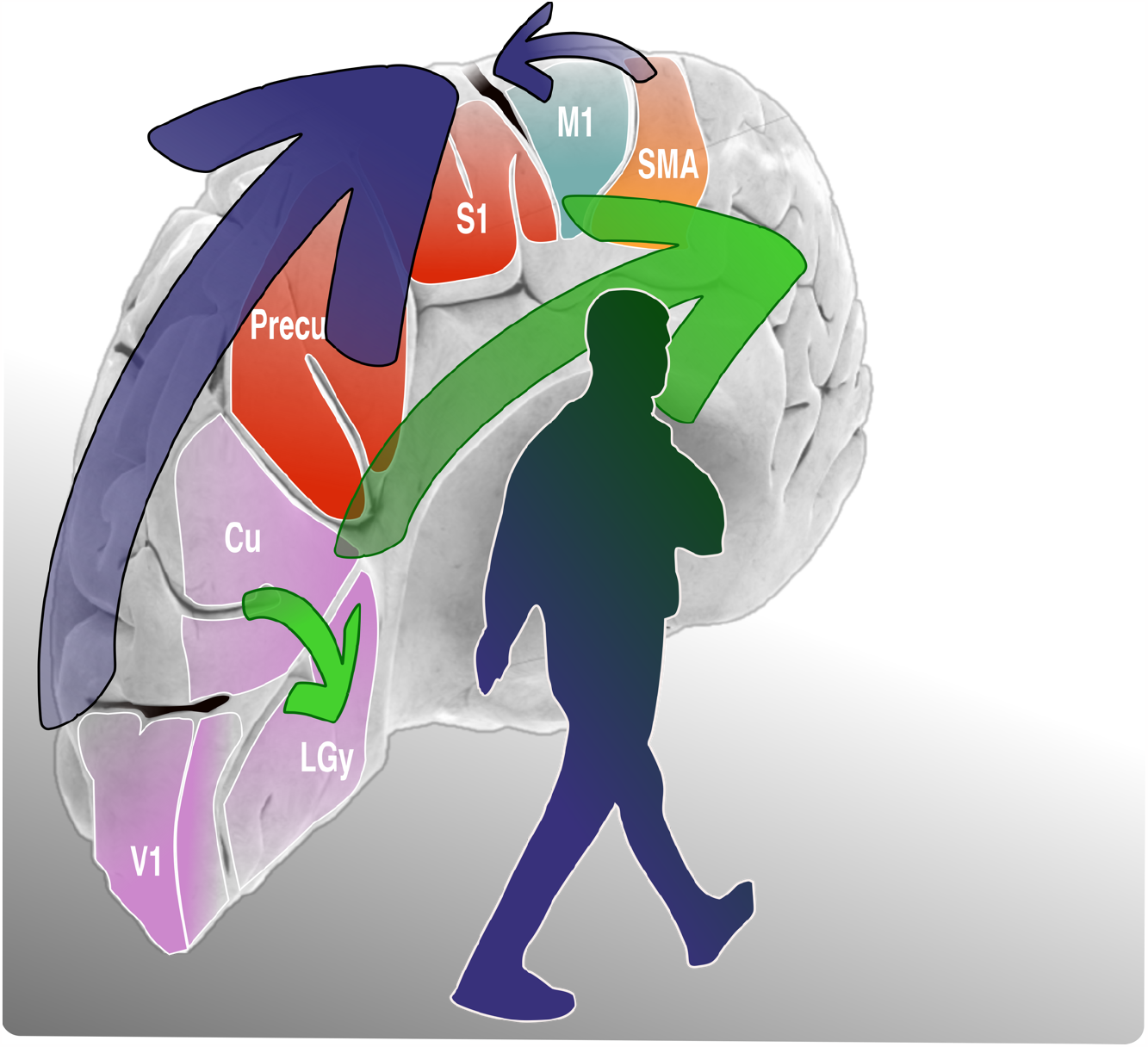
Scheme of the roadblocks and rerouting of locomotion information in PD. In PD, the information flow from visuomotor areas to SMA and from the SMA to sensory areas is impaired (blue arrows), so that a (likely compensatory) rerouting of the locomotor information through the lingual gyrus via the inferior fronto-occipital fasciculus is needed to push forward the locomotor information to sensorimotor brain areas (green arrows). SMA: supplementary motor area; M1: primary motor area (i.e., paracentral gyrus), S1: primary sensory cortex (i.e., postcentral and parietal superior gyrus); Precu: precuneus; Cu: cuneus; LGy: lingual gyrus; V1: calcarine cortex.

### Deficient SMA functional connectivity impairs locomotor control

We found a strong reduction of SMA functional connectivity to the main sensory areas (i.e., paracentral lobule, postcentral lobules and precuneus) during walking as compared to resting in unmedicated PD patients (Figure 4). Reduced information flow between SMA and precuneus lead to hypokinetic gait (Figure 5). An interaction between these areas is essential as feedforward locomotor control originating from the SMA needs to be integrated with somatosensory information to update the body schema and to maintain postural control ^35,36^. Several studies reported abnormalities in sensorimotor integration in subjects with PD ^5^/10/24 11:33:00 AM with evidence for a primary SMA dysfunction in parkinsonian patients with gait disturbances^37,38^. In particular, functional imaging studies showed that the SMA is hypoactive in PD and this correlates with bradykinesia ^39^ impaired generation of anticipatory postural adjustments (APAs) ^40^ and episodes of gait freezing ^41^. Our findings expand on this topic and highlight a primarily functional impairment of the SMA in locomotion with a decupling with motor and sensory areas during unperturbed, overground walking in unmedicated PD patients.

### Visuomotor processing is critical for gait control

Studies in PD showed that gait performances strongly rely on visual inputs ^42^ and that alterations of visuomotor processing are associated with gait disturbances ^43^. In our work, walking was associated with a primary involvement of visuomotor cortex, but with a reduced information flow from these areas to both SMA, motor and sensory cortices (Figure 4). The correlation of functional connectivity patterns of visuomotor areas with gait performances yielded opposing results. Specifically, hypokinetic gait was related to poor information flow from calcarine to lingual gyrus and high information from calcarine to cuneus (Figure 5). Functional imaging studies showed that the lingual gyrus is preferentially involved in direction and discrimination of motion ^44,45^, whereas the cuneus is primarily engaged in basic visual processing (i.e., spatial orientation, frequency, and speed). While a direct engagement in locomotion control of these two areas still need to be proven, preliminary evidence showed that functional connectivity across the bilateral lingual gyrus, calcarine, cuneus, precuneus, and SMA is reduced in PD patients with *freezing of gait* as compared to patients without gait impairment ^46^. Furthermore, PD-related degeneration of neurons in the lingual gyrus and the cuneus have been associated with the occurrence of gait disturbances ^47^.

### Roadblocks and rerouting: the long journey of locomotor information

Altogether our findings detail a roadmap of locomotor information processing in PD and highlight negative functional connectivity changes that may act as roadblock for sensory and visuomotor information processes leading to hypokinetic gait.

The framework of cortical locomotor control starts with sensory inputs (i.e., visual, sensory and proprioceptive information) that generate an internal schema of body and space, which is transmitted to the SMA to be integrated into feedforward locomotor programs. We showed that this process is altered in subjects with PD and highlighted two main functional roadblocks. The first is the impaired information flow from visuomotor areas to SMA that hampers visual integration processing relevant for the construction of APAs (Figure 6). The second roadblock lays in reduced information flow form the SMA to sensory areas that further impairs APAs actualization (Figure 6).

In this scenario, we suggest that an information flow reinforced by the lingual gyrus may act as compensatory strategy to allow the forward processing of visual and locomotor information. This functional rerouting is supported by the articulated anatomical connections of visuomotor areas with frontal areas known as inferior fronto-occipital fasciculus (IFOF) ^45^. The IFOF connects the cuneus and lingual gyrus to the frontal cortex with fibers from the lingual gyrus terminating mostly in the inferior frontal gyrus and fibers of from the cuneus terminating mostly in the superior frontal gyrus. This wide distribution of cortical terminations would provide additional multimodal information processing fostering locomotion ^48,49^.

### Limitations

This study has limitations. First, the relatively small sample size, which mainly derives from the demanding and multimodal approach with multiple brain molecular imaging studies, neurophysiological and kinematic recordings in meds-off state. This complex assessment is not feasible in advanced stages of PD. However, our sample is comparable with other studies on this topic ^10^ with the additional advantage of leveraging brain metabolic imaging findings for neurophysiological analyses. With this regard, we applied well-established and validated electrophysiological metrics ^26,50^. Second, we limited our EEG analysis to cortical areas and cannot comment on the impact of basal ganglia, thalamus and cerebellum on our findings. Last, the lack of a control group limits the generalization of the findings of this study, which however specifically targeted PD patients. We also cannot comment on the impact of different therapeutic strategies since we limited for now our recordings to unmedicated patients.

## Conclusion

This study investigated the functional activity of the cortical locomotor network in unmedicated PD patients during unperturbed overground walking and support a role for visuomotor integration processes in the rescuing of hypokinetic gait in PD. Further studies are needed to assess the impact of dopaminergic and neuromodulation treatments on these functional connectivity pathways.

## Data Availability

All data produced in the present study are available upon reasonable request to the authors.

## Acknowledgements

The authors would like to thank J. Brumberg for his support with molecular imaging data acquisition, Prof. C. Matthies, Dr. P. Fricke and Dr. R. Nickl for the neurosurgical management of the patients and Prof. J. Volkmann for the valuable comments on the results.

## Author contribution

I.U.I. and A.B. contributed with the conception and design of the study, F.P., C.P., N.G.P., G.M., S.H. and I.U.I. contributed with acquisition and analysis of data. F.P., C.P., N.G.P., S.H. and I.U.I. contributed also with drafting a significant portion of the manuscript or figures.

## Potential conflict of interest

F.P., C.P., N.G.P., G.M., A.B. S.H and I.U.I. declare no conflicts of interest related to this study.

## Data Availability Statement

Due to strict privacy law, data are available upon personal request. Inquires can be sent to the corresponding author (Ioannis U. Isaias, University Hospital Würzburg, Department of Neurology, Josef-Schneider-Straße 11, 97080 Würzburg).

## Funding

The study was sponsored by the Deutsche Forschungsgemeinschaft (DFG, German Research Foundation) - Project-ID 424778381-TRR 295, by the European Union - Next Generation EU - NRRP M6C2 - Investment 2.1 Enhancement and strengthening of biomedical research in the NHS and by the Fondazione Grigioni per il Morbo di Parkinson.

SH and FP were supported by the European Research Council (ERC) under the European Union’s Horizon 2020 research and innovation program (Grant agreement No. 758985).

IUI was supported by the New York University School of Medicine and The Marlene and Paolo Fresco Institute for Parkinson’s and Movement Disorders, which was made possible with support from Marlene and Paolo Fresco. CP was supported by a grant from the Fondazione Europea di Ricerca Biomedica (FERB Onlus).

## Notes

### Competing Interest Statement

The authors have declared no competing interest.

### Author Declarations

Ethics Committee of the University of Wuerzburg gave ethical approval for this work.

### Summary of Updates

An error in the information about the authors has been corrected.

