## Supplementary for "Cortical networks of parkinsonian gait: a metabolic and functional connectivity study"

**Supplementary Material**

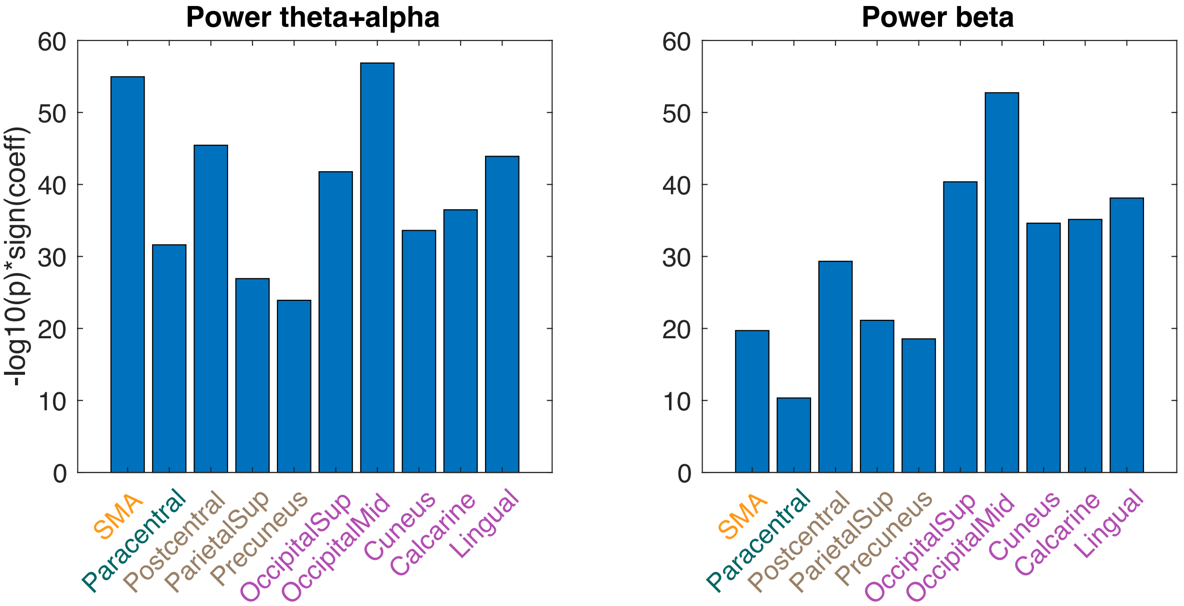


**Figure S1. Comparison of EEG power between resting and walking conditions in the theta-alpha and beta bands.** *We show $log_{10}$ p-values, thus values below -2 and above 2 would indicate significance at alpha level 0.01. Signed $log_{10}$ p-values that were not significant after FDR correction are set to zero.*

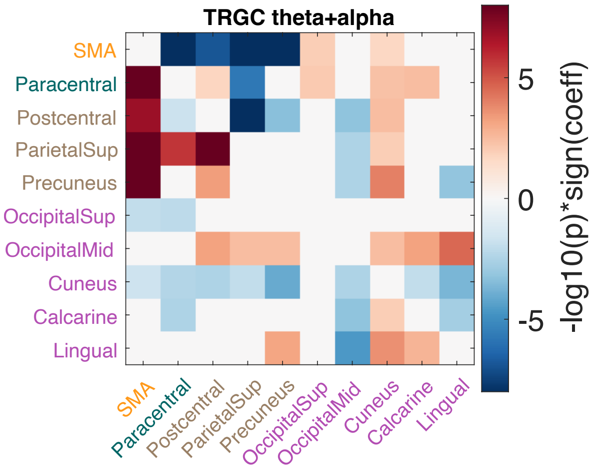


**Figure S2. Comparison of EEG markers between resting and walking conditions in the theta-alpha band.** *Changes in directed functional connectivity were estimated by time-reversed Granger causality (TRGC). Red colors either indicate more information flow from the ROI on the Y-axis to the ROI on the X-axis in walking compared to resting, or less information flow from the ROI shown on the X-axis to the ROI shown on the Y-axis. We show $log_{10}$ p-values, thus values below -2 and above 2 would indicate significance at alpha level 0.01. Signed $log_{10}$ p-values that were not significant after FDR correction are set to zero.*
